# Prevalence of Depression among Senior Secondary School Students in Enugu Metropolis

**DOI:** 10.1101/2023.10.12.23296984

**Authors:** Nzubechukwu Gift Okeke, Johncross Ogbonna Okenyi, Eberechi Cyprian Ohanu, Chidera Okeke

**Author notes:** **SUPERVISOR**. **PROF. AGUWA EMMANUEL**. **FEBRUARY, 2021**. **APPROVAL**. This project on “**PREVALENCE OF DEPRESSION AMONG SENIOR SECONDARY SCHOOL STUDENTS IN ENUGU METROPOLIS**” has been approved by the department of community medicine, College of Medicine, University of Nigeria Enugu Campus. **Prof. E.N. Aguwa**. **(Project Supervisor)**. **Prof. E. Nwobi**. **(Head of Department)**. **DEDICATION:** We dedicate this work first to God Almighty, our beloved families and to every adolescent living with depression.

## Abstract

**Background:** Depression is defined as a state of low mood and aversion to activity. A mental health disorder characterized by persistent low mood or loss of interest in activities causing significant impairment in daily life. Depression amongst secondary school students is an issue that is often ignored. It is frequently unrecognized when present in adolescents who make up the population of secondary school students (age 10-19 years). In our environment, studies have reported that 21.2% of Nigerian adolescents are suffering from mild to moderate depression and depression when left unchecked and untreated, takes a toll on the quality of life of individuals affected. This is why this study was conducted to determine the prevalence of depression among senior secondary school students in Enugu metropolis, to create proper awareness and sensitize people involved and the public on this menace.

**Method:** A descriptive cross-sectional study was carried out with a sample size of 270 secondary school students across 10 secondary schools in Enugu metropolis. The population of 270 students was sampled using simple random sampling. All met the inclusion criteria. A self-administered semi-structured questionnaire was used for data collection. Data was analyzed using statistical package for social sciences (SPSS) version 22. Data was presented using tables.

**Result:** The study had a 100% response rate. Results showed the prevalence of mild depression amongst the respondents as 58.1%, the prevalence of moderate depression amongst the respondents as 31.1% while the prevalence of severe depression was 10.7%. The study found the prevalence of depression to be higher in older adolescents and females. There was also a significant relationship between satisfaction with academic performance and depression.

**Conclusion:** There was significant level of prevalence of depression among senior secondary school students in Enugu metropolis, notably among the females, older adolescents and those who were not satisfied with their academic performance.

**Recommendations:** Depression should be seen as what it is, a disease and awareness should be carried out to adequately sensitize the involved population on early recognition of signs and symptoms of depression, appropriate support and therapy and subsequent rehabilitation of depressed individuals.

## CHAPTER ONE

### 1.1 Background

Depression amongst secondary school students is an issue that is often ignored. It is an uncommon condition and frequently unrecognized when present in adolescents who make up the population of secondary school students. Depression is defined as a state of low mood and aversion to activity. A mental health disorder characterized by persistent low mood or loss of interest in activities causing significant impairment in daily life.

Depression is a serious mental disorder which can often have an impact on social functioning, relationship with family and peers and academic performance in adolescents. Depression amongst this population seems to be evident at earlier ages and may occur with comorbid psychiatric disorders, increased risk for suicide, substance abuse and behavioral problems.

As unrecognized as depression is among the adult population, it is even more so among adolescents. This study is aimed at finding the prevalence of depression among secondary school students in a specified area. Depression is a real illness that impacts on the brain. Depressive symptoms present in adolescents are often overlooked and attributed to normal life stress associated with that stage of life and other times misdiagnosed as substance abuse disorders and this act of neglect can have very tragic consequences considering that adolescent who experience depression at an early stage often struggle with it during adulthood.

Globally, the prevalence of mental health problems is 10% −20% among children and adolescents ^[1]^. Studies have estimated that depression affects 8.3% of older adolescents in the United States. In addition, it is noted that on any single day, about 8% of adolescents meet the criteria for major depression. In the long run, the numbers have increased; the rates of depression are as high as 28% for adolescents in primary care settings.

To further bring this home, studies have reported that 21.2% of Nigerian adolescents are suffering from mild to moderate depression, while a study done by Fatrigun and Kumapayi showed that 5.7% are severely depressed ^[2]^. The consequences of not addressing the issue of mental health conditions especially depression in secondary school students extend to adulthood, impairing both physical and mental health and thus limiting opportunities to lead fulfilling lives as adults.

### 1.2 Problem Statement

The age group found in the secondary school population is the age of adolescence which is crucial period for developing and maintaining social and emotional habits important for mental well-being. These include adopting healthy sleep patterns, taking regular exercise, developing and coping with circumstances around them, problem solving, acquiring interpersonal skills and learning how to manage emotions.

Depression in this age group alters natural progression of these growth processes and has dire consequences on the sufferer, the family, and the society at large. It becomes the starting point of the many problems the individual is predisposed to. These problems will be mentioned briefly below.

Emotional disorders commonly emerge during adolescence. In addition to depression, adolescents easily develop emotional disorders and experience excessive irritability, frustration and anger which culminate in dysfunctional relationships and markedly reduced productivity especially with academic performance.

Many risk taking behaviours such as sexual risk taking start during adolescence and most victims are driven into these vices by episodes of depression that were unattended.

Perpetration of violence is also a risk taking behaviour that is triggered by depression and can increase the likelihood of low educational attainment, injury, involvement with crime and death.

The use of tobacco and cannabis is often associated with adolescents and not so farfetched in their of them suffering depression and seeking means of escape from their predicament An estimated 62,000 adolescents died in 2016 as a result of self-harm. Suicide is the third leading cause of death in older adolescents (15-19 years) and is often linked to depressive symptoms.

### 1.3 Justification

There is need for this study because there is insufficient data and information pertaining the mental health of adolescents in secondary schools in Nigeria as previous studies which varied greatly in their methodology have focused more on adults than on adolescents. To make matters worse there is virtually little to none to highlight the possible factors influencing depression in adolescents in this region. Consequently, it is not surprising that little has been mentioned regarding the services available to assist these adolescents and their families who invariably form an integral part of the social support system.

#### Aim

To assess the prevalence, levels of depression and factors associated with depression among secondary school adolescents in Enugu metropolis

### 1.4 Research questions

a. What is the prevalence of depression among senior secondary school students in Enugu Metropolis?
b. What is the level of depression among senior secondary school students in Enugu metropolis?
c. What are the factors that influence depression among senior secondary school students in Enugu metropolis?
d. What are the health risk behaviors associated with depression among senior secondary school students in Enugu metropolis.

### 1.5 Objectives

1. To identify prevalence of depression among senior secondary school students in Enugu metropolis.
2. To identify the levels/scales of depression among senior secondary students in Enugu metropolis
3. To determine the socio-demographic factors associated with depression among senior secondary students in Enugu metropolis
4. To determine the health risk behaviors associated with depression among senior secondary school students in Enugu metropolis.

## CHAPTER TWO

### 2.1 Overview of Depression

Depression is a common illness worldwide, with more than 264 million people affected ^[23]^. Depression is different from usual mood fluctuations and short-lived emotional responses to challenges in everyday life. Especially when long-lasting and with moderate or severe intensity, depression may become a serious health condition. It can cause the affected person to suffer greatly and function poorly at work, at school and in the family. At its worst, depression can lead to suicide. Close to 800 000 people die due to suicide every year. Suicide is the second leading cause of death in 15-29-year-olds ^[24]^

It is a mental health disorder characterized by persistently depressed mood or loss of interest in activities, causing significant impairment in daily life. Possible causes include a combination of biological, psychological and social sources of distress. Increasingly, research suggests that these factors may cause changes in brain function, including altered activity of certain neural circuits in the brain. The most common symptom is a low mood that can sometimes appear as irritability. Often the person with depression is not being able to enjoy activities that he or she normally enjoys. Many people with depression also have anxiety. They may worry more than average about their physical health. They may have excessive conflict in their relationships or function poorly at work. Sexual functioning may be a problem. People with depression are also at more risk for abusing alcohol or other substances.

Depression probably involves changes in the areas of the brain that control mood. The nerve cells may be functioning poorly in certain regions of the brain. Altered communication between nerve cells or nerve circuits can make it harder for a person’s brain to regulate his or her mood^[25]^ Hormone changes may also negatively affect mood. An individual’s life experiences can affect these biological processes. And a person’s genetic makeup influences how vulnerable he or she is to experiencing depression. An episode of depression can be triggered by a stressful life event. But in many cases, depression does not appear to be related to a specific event. A major depressive episode may occur within the first two to three months after giving birth to a baby. In that case, it may be called major depressive disorder with peripartum onset. Most people refer to it as postpartum depression. ^[25]^ Depression is diagnosed in women twice as often as in men. People who have a family member with major depression are more likely to develop depression.^[25]^

#### Symptoms

A depressed person may gain or lose weight, eat more or less than usual, have difficulty concentrating, and have trouble sleeping or sleep more than usual. He or she may feel tired and have no energy for work or play. Small burdens or obstacles may appear impossible to manage. The person can appear slowed down, or agitated and restless. The symptoms can be quite noticeable to others. A particularly painful symptom of this illness is an unshakable feeling of worthlessness and guilt. The person may feel guilty about a specific life experience, or may feel general guilt not related to anything in particular.

Although depression may occur only once during one’s life, people typically have multiple episodes. During these episodes, symptoms occur most of the day, nearly every day and may include: feelings of sadness, tearfulness, emptiness or hopelessness, angry outbursts, irritability or frustration, even over small matters. Loss of interest or pleasure in most or all normal activities, such as sex, hobbies or sports, Sleep disturbances, including insomnia or sleeping too much, tiredness and lack of energy, so even small tasks take extra effort, reduced appetite and weight loss or increased cravings for food and weight gain, anxiety, agitation or restlessness, slowed thinking, speaking or body movements, feelings of worthlessness or guilt, fixating on past failures or self-blame, trouble thinking, concentrating, making decisions and remembering things, frequent or recurrent thoughts of death, suicidal thoughts, suicide attempts or suicide unexplained physical problems, such as back pain or headaches.

For many people with depression, symptoms usually are severe enough to cause noticeable problems in day-to-day activities, such as work, school, social activities or relationships with others. Some people may feel generally miserable or unhappy without really knowing why.

#### Depression symptoms in children and teens

Common signs and symptoms of depression in children and teenagers are similar to those of adults, but there can be some differences. In younger children, symptoms of depression may include sadness, irritability, clinginess, worry, aches and pains, refusing to go to school, or being underweight. In teens, symptoms may include sadness, irritability, feeling negative and worthless, anger, poor performance or poor attendance at school, feeling misunderstood and extremely sensitive, using recreational drugs or alcohol, eating or sleeping too much, self-harm, loss of interest in normal activities, and avoidance of social interaction.

The global prevalence of depression and depressive symptoms has been increasing in recent decades. The lifetime prevalence of depression ranges from 20% to 25% in women and 7% to 12% in men. ^[26]^ Although there are known, effective treatments for mental disorders, between 76% and 85% of people in low- and middle-income countries receive no treatment for their disorder.^[27]^ Barriers to effective care include a lack of resources, lack of trained health-care providers and social stigma associated with mental disorders. Another barrier to effective care is inaccurate assessment. In countries of all income levels, people who are depressed are often not correctly diagnosed, and others who do not have the disorder are too often misdiagnosed and prescribed antidepressants.

The burden of depression and other mental health conditions is on the rise globally. A World Health Assembly resolution passed in May 2013 has called for a comprehensive, coordinated response to mental disorders at the country level ^[28]^

### 2.2 : Types of depression

The main types of depression include:

Major depression

It occurs when feelings of sadness, loss, anger, or frustration interfere with daily life for weeks or longer periods of time. Loss of interest or pleasure in activities, weight loss or gain, insomnia, restlessness or agitation, Feeling worthless or guilty, trouble concentrating or making decisions suicidal ideations.

Persistent depressive disorder

This is a depressed mood that lasts 2 years. Over that length of time, one may have periods of major depression, and with times when the symptoms are milder one may have symptoms such as:

Change in appetite (not eating enough or overeating), sleep too much or too little lack of energy, or fatigue, low self-esteem, trouble concentrating or making decisions, feeling of hopelessness. One may be treated with psychotherapy, medication, or a combination of the two.

**Other common forms of depression include:**

Postpartum depression

Many women feel somewhat down after having a baby. However, true postpartum depression is more severe and includes the symptoms of major depression.

Premenstrual dysphoric disorder (PMDD)

Symptoms of depression occur 1 week before menstrual period and disappear after menstruation. Women with PMDD have depression and other symptoms at the start of their period. Besides feeling depressed, there may also be:

Mood swings, Irritability, Anxiety, Trouble concentrating, Fatigue, Change in appetite or sleep habits, Feelings of being overwhelmed

Antidepressant medication or sometimes oral contraceptives can treat PMDD.

Seasonal affective disorder (SAD)

This occurs most often during fall and winter, and disappears during spring and summer. It is most likely due to a lack of sunlight.

Seasonal affective disorder is a period of major depression that most often happens during the winter months, when the days grow short when people get less and less sunlight. It typically goes away in the spring and summer.

Major depression with psychotic features

This occurs when a person has depression and loss of touch with reality (psychosis).Also known as “Psychotic Depression”.

People with psychotic depression have the symptoms of major depression along with “psychotic” symptoms, such as: Hallucinations (seeing or hearing things that aren’t there), Delusions (false beliefs), and paranoia (wrongly believing that others are trying to harm you)

Bipolar disorder

This occurs when depression alternates with mania (formerly called manic depression). Bipolar disorder has depression as one of its symptoms, but it is a different type of mental illness.

#### Atypical Depression

Atypical depression is a subtype of major depression that involves several specific symptoms, including increased appetite or weight gain, sleepiness or excessive sleep, marked fatigue or weakness, moods that are strongly reactive to environmental circumstances, and feeling extremely sensitivity to rejection or being oversensitive to criticism. It is considered to be a “specifier” that describes a pattern of depressive symptoms. If you have atypical depression, a positive event can temporarily improve your mood. ^[29]^

### 2.3 Prevalence of depression in Sub Saharan Africa

Mental health problems appear to be increasing in importance in Africa. Between 2000 and 2015 the continent’s population grew by 49%, yet the number of years lost to disability as a result of mental health disorders increased by 52%. ^[4]^

The prevalence of Major depressive disorder remains largely unknown in many sub-Saharan African countries. Studies carried out to date have been single-country studies, almost exclusively in Anglophone countries, rarely conducted at national level, and have produced a wide range of prevalence estimates. Two recent studies that estimated the 12-month prevalence for major depressive episode found low or moderate rates, including a survey conducted by the World Health Organization (WHO) in Nigeria which found a prevalence of 1 %.^[5]^

It is also important to note that rapid urbanization in countries in sub Saharan Africa comes with a lot of social changes which take a toll on individual’s mental health. Poverty, Unemployment, Stress and violence levels are indices that greatly contribute to one’s mental health status.

### 2.4 : Prevalence and Predictors of depression in adolescents and secondary school students. Prevalence

Depression among adolescents has been of recent increasingly recognized as a public health problem all over the world. Although it was rightly noted in the background of this study that depression among adolescents who make up the population of secondary schools is often overlooked and most times misdiagnosed, more and more studies have been done over time on epidemiology of depression among the said group.

More so, studies on prevalence and independent indices that affect rates of depression in developing countries especially in sub Saharan Africa should be greatly encouraged.

In Nigeria, as in most developing countries, there is a need for more research on the epidemiology of adolescent depression as this will guide prevention, diagnosis and treatment. ^[6]^

The prevalence of depression among secondary school students rises substantially throughout teenage years which could be due to pronounced biological and social changes which they inevitably go through at home, school, and other parts of the society they belong in.

Diagnostic Interview Schedule for Children (DISC) reported a prevalence rate of probable depression in Adolescents to be 12.1%. This prevalence is slightly lower than 16.3% obtained in a study done by Oderinde et al. ^[6]^ The variation in the finding from Omigbodun et al. ^[9]^ study compared to the current study may be due to varying methodological factors.

A more recent study by Fatiregun and Kumapayi using Patient Health Questionnaire modified for adolescents reported the prevalence of depressive symptoms to be 21.2% among school adolescents in a rural district of Egbeda Local Government Area of Oyo State ^[10]^

In other parts of developing world such as India, Nagendra et al.^[11]^ using the Becks Depression Inventory reported a one month prevalence rate of depressive symptoms to be 57.7% in a sample of 3141 adolescent students within the age range 15 - 19 years among public and private secondary school students.

The study done by Oderinde also found that the proportion of older adolescents in the age range 16 - 19 years who were found to be depressed was higher than the proportion of younger adolescents in the age range 10 - 12 years who were found to be depressed.^[6]^ Possible reasons for this could be, societal expectations from older adolescents such as better academic performance, better conduct than the younger ones, hence they are more likely to experience more stressors than the younger adolescents.^[12]^ Hormonally-linked heightened stress sensitivity in the older adolescents has been associated with higher prevalence estimates of depression among the older adolescents. This is because hormonal changes produce behavioral and neural signs of depression by sensitizing the brain to the harmful effects of stress. ^[13]^

In Nigeria, Omigbodun et al. reported 12.6% as prevalence of probable depression among adolescents in rural South West Nigeria and found experiencing traumatic events as one of the predictors of depression especially when the event directly affected the youth as in sexual assault or physical abuse.^[8]^

#### Predictors

Studies have been done to show relationship between some named factors and depression in adolescents and secondary school students. These factors include

**Gender**: This was among the most significant predictors of depression in a study done by Al-Kaabi et al ^[14]^ as it has been found that females were more likely to have depression than males and this comes in agreement with many studies and the fact that females are more susceptible to depression may be explained by the theory that the biological and physical makeup of females automatically puts them more at risk of developing depression, as from puberty onwards, fluctuating hormone levels affects their body both physically and emotionally.^[15,16]^

In addition to biological factors, psychological factors may play a substantial role in the predisposition of females to depression. Although times are indeed changing, young women are still nevertheless being raised to be subordinate to men in the sense that they are still schooled to be careers and nurturers. They also tend to be more affected by the environment around them, and strive for perfection both physically and otherwise. ^[14]^

This predefined social role, both increases the pressure, which they place on themselves to please others and increases the likelihood that they will be subjected to some form of abuse during their lifetime ^[14]^ hence increasing the likelihood of the occurrence of depression in this gender.

**Obesity**: This variable expressed by BMI was found to be significantly related to depression in the study done by Al–Kaabi et al. ^[14]^ and this is in agreement with many studies, but the evidence for the direct causal pathway from obesity to depression is not substantial. Obesity might not directly cause depression in adolescents, but other pathways and experiences may lead to depression indirectly as stressful life events such as peer victimization and weight-based teasing might biologically predispose youth to depression and may be a factor that leads to depression in obese youth.

Further research studies exploring these factors in youth will increase our understanding of obesity/depression associations and might then be a venue for intervention studies. The importance of recognizing these pathways and factors are to know when to intervene to prevent depression in obese adolescents ^[14]^

**Family Conflict**: It is known that verbal and physical quarrels of parents have bad effects on the mental health of students. The current study done by Al-Kaabi showed a significant relationship between Participants whom were having conflicts in their families and depression, these factors were among the most significant predictor of depression in the logistic regression and this comes in agreement with many studies, where these findings reflect the conflict within the family environment which leads to depression.^[14]^

**Parent-Adolescent relationships:** If characterized by warmth and involvement may protect youth from adjustment problems. A study done by Al-Kaabi et al showed that bad relationships with parents were significantly related to depression among adolescents and as those with bad relationships with parents are 9.4 times more likely to have depression, this comes in agreement with many studies conducted in the literature addressing this issue such as Branje et al. ^[17]^

Presence of harmonious relationships can ensure the stability of the family and be a protective factor against life stresses.

#### Socio-economic class

Socioeconomic class was also found to be significantly associated with depression in a study done by Oderinde et al as students from lower socioeconomic status were six times more likely to be depressed than students from the upper socioeconomic status. ^[6]^

This has been observed in other similar studies conducted in other parts of the world where they found that poverty and difficulties in meeting daily necessities may induce a child to compare himself with others and this situation increases child’s tendency to depression ^[18,19]^

#### Polygamous families

The study by Oderinde et al also found that adolescents from polygamous families were almost six times more likely to be depressed than those from monogamous families and this was significantly and independently associated with depression on logistic regression. Previous studies have also linked polygamous family settings to adolescent depression. This might be due to parents in polygamous family settings being unable to fulfill the needs of their growing children as a result of more numbers in the family. Such needs would include food, clothing, education, love, care, emotional support, parental support and financial needs. ^[6]^

Other factors consistent with depression in adolescents and secondary school students in literature reviewed include;

**Parental separation/divorce**; Adolescents from separated/divorced families tend to be more depressed than their same aged peers from intact families. This could be, because such adolescents from divorced home tend to have less intimate relationship with their parents.

**Death of a parent**; this was also found to be significantly associated with adolescent depression in various studies. Previous researchers have reported that adolescents who had experienced parental death were more likely to be depressed than those who had not ^[20,21]^. The reason for this observation is obvious, since such adolescents are often deprived of secure and loving relationships with their parents and these are protective factors that reduce the rate of emotional disorders among adolescents ^[22]^

### 2.5 : Health Risk Behaviours associated with Depression

Risk-taking includes behaviours that have a chance of a desired, beneficial outcome but with the possibility of unwanted, negative consequences. Health risk behaviours take a toll on the individual’s in the long run. Adolescence and young adulthood are characterized by a disproportionate increase in risk-taking behaviour.^[34]^

An increasing number of studies have pointed to an association between depressive disorders and various health risk-related behaviours in various population groups (general population, adolescents, adults, elderly, etc.^[32]^

These health risk behaviours include tobacco use or dependency, alcoholism, self-harm, risky sexual behaviors, insufficient physical or sporting activity, and unhealthy eating habits. ^[32]^

A few studies, which mainly concern adolescents, have sought to determine whether or not depression increases the probability of co-occurrence of several health risk behaviours. These studies suggest that risk-related behaviours tend to cluster or co-occur rather than occurring in isolation. ^[32]^

Adolescence is a vulnerable period which is associated with a heightened risk for the development of depressive disorders. Most risk-behaviors tend to be associated with increased likelihood for the development of depression and are correlated with the severity of depressive symptoms. ^[33]^

## CHAPTER THREE

### METHODOLOGY

#### 3.1. Study Area

Study area is Enugu state. Enugu State is located in the south east geopolitical zone of Nigeria. It shares boundary with Anambra on the west, Abia State on the south, Kogi on the north, and Benue and Ebonyi on the east. Enugu was the headquarters of the former East Central State and Eastern Nigeria ^[30]^.

There are seventeen local government areas (LGAs) in the state with a total population of 3,267,837 (5,590,513 - 2006 estimate) people. The State has three urban centers: Enugu, Nsukka and Oji River. The state capital is a modern city which covers an area of 85 sq. km with a population of about 717,291 people. It has well developed coal mining, commercial, financial and industrial center, with booming economy and vast investment opportunities. The state is predominantly agrarian with yam tubers, palm produce and rice being their main produce. ^[30]^

#### 3.2: Study Population/ Participants

This study was conducted among senior secondary school students in secondary schools in Enugu Metropolis

#### 3.3: Study Design

A descriptive cross-sectional study was carried out among senior secondary school students in selected 10 public and private secondary schools within Enugu Metropolis.

#### 3.4. Sampling Technique

A list of secondary schools in Enugu metropolis was obtained from the state ministry of Education and 10 secondary schools were randomly selected via simple random sampling method. A total of 27 students were selected equally from each school using the simple random sampling technique to make up the required sample size.

From each arm (SS1, SS2 and SS3), using the class list, 9 students each were randomly selected using a simple random sampling method

#### 3.5. Sample size estimation

n = Z^2^P(1-P)/ D^2^

Zα = Confidence level is 95%, Zα = 1.96

P = Prevalence of depression in Nigerian Adolescents = 21.2%^[2]^

D = Margin of error tolerated =5% = 0.05

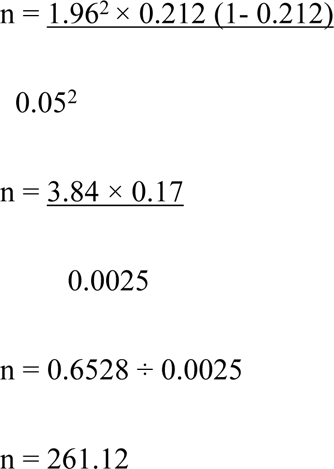

Sample size = 261.12.

Therefore, 270 participants will be included in the study

#### 3.6. Inclusion Criteria

This study included students who are within 14 to 20 years old in senior secondary classes 1-3 in selected secondary schools in Enugu Metropolis and students within the said range who were willing to participate.

#### 3.7. Exclusion Criteria

Students who met the inclusion criteria but were not willing to participate were excluded from the study.

#### 3.8. Sampling Instrument

A semi-structured self-administered questionnaire was used for data collection. It consists of 3 sections. A collected socio demographic data, section B collected data on health risk behaviours, section C collected data on levels of depression using the Beck’s depression tool.

Variables in the questionnaire were adapted from Beck’s depression tool ^[31]^ and Youth Risk Behaviour Surveillance System (YRBSS) ^[35]^

#### 3.9. Data Collection

A quantitative data was collected using a semi structured, self-administered questionnaire. The questionnaire was distributed by members of the research team after written informed consent was obtained.

#### 3.10. Data Analysis

Data was analyzed using Statistical Package for Social Sciences (SPSS) version 23.0, results was displayed in tables and pie charts. Descriptive analysis will be carried out and factors associated with depression will be carried using Chi square test of significance and P value will be set at 0.05.

#### 3.11. Ethical Consideration

##### Ethical Clearance

Ethical clearance for the study was obtained from the Health Research Ethics Review Committee of University of Nigeria Teaching Hospital (UNTH), Ituku-Ozalla.

##### Informed Consent

A written informed consent was obtained from the guardians of minors who were respondents and other respondents, where they were told about the research and that they were free to opt-out of the study at any time without any negative consequences.

##### Confidentiality

Information that was retrieved from respondents was handled with utmost respect. Care was taken to ensure that the participants are kept anonymous during and after the study. And data collected from respondents were treated purely as academic and confidential for the safety, social and psychological well-being of the respondents. Authors did not have access to information that could identify participants during or after data collection.

## CHAPTER FOUR

### RESULTS

**Table 4.1:** Sociodemographic characteristics of respondents.

| *Variable* | *Value* | *Frequency*<br>*(N=270)* | *Percentage*<br>*(%)* |
| --- | --- | --- | --- |
| **Age** | **14 years** | 19 | 7.0 |
|  | **15 years** | 74 | 27.4 |
|  | **16 years** | 75 | 27.8 |
|  | **17 years** | 60 | 22.2 |
|  | **18 years** | 17 | 6.3 |
|  | **19 years** | 13 | 4.8 |
|  | **20 years** | 12 | 4.4 |
| **Ave age: 16.26 yrs** |  |  |  |
| **Std dev: 1.46 yrs** |  |  |  |
| **Class** | **SSS1** | 43 | 15.9 |
|  | **SSS2** | 169 | 62.6 |
|  | **SSS3** | 58 | 21.5 |
| **Sex** | **Male** | 88 | 32.6 |
|  | **Female** | 182 | 67.4 |
| **Tribe** | **Igbo** | 264 | 97.8 |
|  | **Yoruba** | 3 | 1.1 |
|  | **Others** | 3 | 1.1 |
| **State of Origin** | **South-East** | 248 | 91.9 |
|  | **South-South** | 5 | 1.9 |
|  | **South-West** | 11 | 4.1 |
|  | **North** | 6 | 2.2 |
| **Religion** | **Christian** | 268 | 99.3 |
|  | **Islam** | 2 | 0.7 |
| **Father's occupation** | **Business** | 146 | 54.0 |
|  | **Civil servant** | 50 | 18.5 |
|  | **Professional** | 49 | 18.1 |
|  | **No occupation** | 25 | 9.3 |
| **Mother's occupation** | **Business** | 135 | 50.0 |
|  | **Civil servant** | 89 | 33.0 |
|  | **Professional** | 27 | 10.0 |
|  | **No occupation** | 19 | 7.0 |

The average age of the respondents is 16.26 + 1.46 years. Majority of the respondents are within the age of 15 to 17 years. Most of the respondents, 62.6% are from SSS2 class. About 32.6% were males while 67.4% were females. Almost all the respondents are of the Igbo tribe from the South Eastern part of the country and are also Christians. A majority of the respondents parents occupation is mainly business.

**Table 4.2:** Family and Social history of respondents.

| ***Variable*** | ***Yes***<br>***(%)*** | ***No***<br>***(%)*** |
| --- | --- | --- |
| **Are both parents alive?** | 256 (94.8%) | 14 (5.2%) |
| **Are you living with your parents?** | 237 (87.8%) | 33 (12.2%) |
| **Are your parents living together?** | 247 (91.5%) | 23 (8.5%) |
| **Do you have a boyfriend/ girlfriend?** | 95 (35.2%) | 175 (64.8%) |
| **Do you use social media?** | 191 (70.7%) | 79 (29.3%) |
| **Are you satisfied with your academic performance?** | 157 (58.1%) | 113 (41.9%) |
| **Do you receive monthly allowance?** | 62 (23.0%) | 208 (76.7%) |

Both parents are alive and are living together for almost all the respondents. A majority of the respondents live with their parents. From the table, 70.7% of the respondents use social media while only 58.1% are satisfied with their academic performance. Only a few, 23.0% do receive monthly allowances.

**Table 4.3:** Other Family and Social history of respondents.

| *Variable* | *Value* | *Frequency*<br>*(N=270)* | *Percentage*<br>*(%)* |
| --- | --- | --- | --- |
| **What is your birth position** | **1** | 86 | 31.9 |
|  | **2** | 77 | 28.5 |
|  | **3** | 50 | 18.5 |
|  | **4** | 35 | 13.0 |
|  | **5** | 10 | 3.7 |
|  | **6** | 7 | 2.6 |
|  | **7** | 4 | 1.5 |
|  | **9** | 1 | 0.4 |
| **How much time do you spend on social media?** | **0 to 1 hour** | 59 | 21.9 |
|  | **2 hours** | 36 | 13.3 |
|  | **3 hours** | 25 | 9.3 |
|  | **4 to 6 hours** | 18 | 6.7 |
|  | **6 to 12 hours** | 9 | 3.3 |
|  | **The whole day** | 2 | 0.7 |
|  | **No idea** | 121 | 44.8 |
|  | **Don't use social media** | 79 | 29.3 |
| **How much allowance do you receive monthly?** | **N0 to N1000** | 34 | 12.6 |
|  | **N1000 to N5000** | 16 | 5.9 |
|  | **N5000 to N10000** | 9 | 3.3 |
|  | **N10000 to N25000** | 2 | 0.7 |
|  | **>N25000** | 1 | 0.4 |
|  | **I don't receive allowance** | 208 | 76.7 |

**Table 4.4:** Health risk behavior assessment of respondents.

| *Variable* | **Yes [1]**<br>**(%)** | **No [0]**<br>**(%)** |
| --- | --- | --- |
| **Have you ever tried smoking?** | 21 (7.8%) | 249 (92.2%) |
| **Have you ever taken alcoholic drinks?** | 178 (65.9%) | 92 (34.1%) |
| **Have you used marijuana or weed?** | 7 (2.6%) | 263 (97.4%) |
| **Are you sexually active?** | 17 (6.3%) | 253 (93.7%) |
| **Have you been pregnant or gotten someone pregnant?** | 4 (1.5%) | 266 (98.5%) |
| **Do you think you are physically unfit?** | 43 (15.9%) | 227 (84.1%) |
| **Do you want to lose or gain weight?** | 62 (23.0%) | 208 (77.0%) |
| **Have you carried a weapon (gun or knife) to school?** | 6 (2.2%) | 264 (97.8%) |
| **Have you fought in the past 6 months?** | 61 (22.6%) | 209 (77.4%) |
| **Have you ever thought of killing yourself?** | 55 (20.4%) | 215 (79.6%) |

The table above assesses the health risk behaviors of the respondents. Questions affirmed by the respondents gets a score of 1. The total score from each respondent was used to derive the table below.

**Table 4.5:** Health risk behavior score of respondents.

| *Variable* | *Frequency*<br>*(n=270)* | *Percentage*<br>*(%)* |
| --- | --- | --- |
| **No risk (0)** | 8 | 3.0 |
| **Low risk (1 to 2)** | 174 | 64.4 |
| **High risk ( >2)** | 88 | 32.6 |
| **Min score amongst respondents: 0** |  |  |
| **Max score amongst respondents: 6** |  |  |
| **Mean score: 2.22 Std. dev: 1.19** |  |  |

Respondents who scored 0 were classified as no risk while those who score 1 to 2 as low risk and those who scored more than 2 as high risk. Most of the respondents (64.4%) had low risk. The mean score was 2.22 + 1.19.

**Table 4.6a:** Assessment of the level of depression amongst respondents.

| *Variable* | *Value* | *Frequency*<br>*(N=270)* | *Percentage*<br>*(%)* |
| --- | --- | --- | --- |
| 1 | 0 I do not feel sad. | 154 | 57.0 |
|  | 1 I feel sad | 91 | 33.7 |
|  | 2 I am sad all the time and I can't snap out of it. | 11 | 4.1 |
|  | 3 I am so sad and unhappy that I can't stand it. | 12 | 5.2 |
| 2 | 0 I am not particularly discouraged about the future. | 236 | 87.4 |
|  | 1 I feel discouraged about the future | 19 | 7.0 |
|  | 2 I feel I have nothing to look forward to. | 9 | 3.3 |
|  | 3 I feel the future is hopeless and that things cannot improve. | 6 | 2.2 |
| 3 | 0 I do not feel like a failure | 223 | 82.6 |
|  | 1 I feel I have failed more than the average person. | 27 | 10.0 |
|  | 2 As I look back on my life, all I can see is a lot of failures | 15 | 5.6 |
|  | 3 I feel I am a complete failure as a person. | 5 | 1.9 |
| 4 | 0 I get as much satisfaction out of things as I used to | 190 | 70.4 |
|  | 1 I don't enjoy things the way I used to. | 56 | 20.7 |
|  | 2 I don't get real satisfaction out of anything anymore | 13 | 4.8 |
|  | 3 I am dissatisfied or bored with everything. | 11 | 4.1 |
| 5 | 0 I don't feel particularly guilty | 140 | 51.9 |
|  | 1 I feel guilty a good part of the time. | 61 | 22.6 |
|  | 2 I feel quite guilty most of the time | 62 | 23.0 |
|  | 3 I feel guilty all of the time. | 7 | 2.6 |
| 6 | 0 I don't feel I am being punished. | 143 | 53.0 |
|  | 1 I feel I may be punished. | 70 | 25.9 |
|  | 2 I expect to be punished. | 19 | 7.0 |
|  | 3 I feel I am being punished. | 38 | 14.1 |
| 7 | 0 I don't feel disappointed in myself. | 220 | 81.5 |
|  | 1 I am disappointed in myself. | 33 | 12.2 |
|  | 2 I am disgusted with myself. | 10 | 3.7 |
|  | 3 I hate myself. | 7 | 2.6 |
| 8 | 0 I don't feel I am any worse than anybody else. | 170 | 63.0 |
|  | 1 I am critical of myself for my weaknesses or mistakes. | 33 | 12.2 |
|  | 2 I blame myself all the time for my faults. | 43 | 15.9 |
|  | 3 I blame myself for everything bad that happens. | 24 | 8.9 |
| 9 | 0 I don't cry any more than usual. | 178 | 65.9 |
|  | 1 I cry more now than I used to. | 29 | 10.7 |
|  | 2 I cry all the time now. | 15 | 5.6 |
|  | 3 I used to be able to cry, but now I can't cry even though I want to | 48 | 17.8 |
| 10 | 0 I am no more irritated by things than I ever was. | 171 | 63.3 |
|  | 1 I am slightly more irritated now than usual. | 44 | 16.3 |
|  | 2 I am quite annoyed or irritated a good deal of the time. | 40 | 14.8 |
|  | 3 I feel irritated all the time. | 15 | 5.6 |

**Table 4.6b:** Assessment of the level of depression amongst respondents.

| *Variable* | *Value* | *Frequency*<br>*(N=270)* | *Percentage*<br>*(%)* |
| --- | --- | --- | --- |
| 11 | 0 I have not lost interest in other people. | 171 | 63.3 |
|  | 1 I am less interested in other people than I used to be. | 60 | 22.2 |
|  | 2 I have lost most of my interest in other people. | 27 | 10.0 |
|  | 3 I have lost all of my interest in other people. | 12 | 4.4 |
| 12 | 0 I make decisions about as well as I ever could. | 195 | 72.2 |
|  | 1 I put off making decisions more than I used to. | 28 | 10.4 |
|  | 2 I have greater difficulty in making decisions more than I used to. | 43 | 15.9 |
|  | 3 I can't make decisions at all anymore. | 4 | 1.5 |
| 13 | 0 I don't feel that I look any worse than I used to. | 215 | 79.6 |
|  | 1 I am worried that I am looking old or unattractive. | 16 | 5.9 |
|  | 2 I feel there are changes in me that make me look unattractive | 21 | 7.8 |
|  | 3 I believe that I look ugly. | 18 | 6.7 |
| 14 | 0 I can work about as well as before. | 185 | 68.5 |
|  | 1 It takes an extra effort to get started at doing something. | 46 | 17.0 |
|  | 2 I have to push myself very hard to do anything. | 33 | 12.2 |
|  | 3 I can't do any work at all. | 6 | 2.2 |
| 15 | 0 I can sleep as well as usual. | 176 | 65.2 |
|  | 1 I don't sleep as well as I used to. | 60 | 22.2 |
|  | 2 I wake up 1-2 hours earlier than usual and find it hard to get back to sleep. | 23 | 8.5 |
|  | 3 I wake up several hours earlier than I used to and cannot get back to sleep. | 11 | 4.1 |
| 16 | 0 I don't get more tired than usual. | 171 | 63.3 |
|  | 1 I get tired more easily than I used to. | 76 | 28.1 |
|  | 2 I get tired from doing almost anything. | 16 | 5.9 |
|  | 3 I am too tired to do anything. | 7 | 2.6 |
| 17 | 0 My appetite is no worse than usual. | 172 | 63.7 |
|  | 1 My appetite is not as good as it used to be. | 68 | 25.2 |
|  | 2 My appetite is much worse now. | 25 | 9.3 |
|  | 3 I have no appetite at all anymore. | 5 | 1.9 |
| 18 | 0 I haven't lost much weight, if any, lately. | 212 | 78.5 |
|  | 1 I have lost more than five pounds. | 50 | 18.5 |
|  | 2 I have lost more than ten pounds. | 5 | 1.9 |
|  | 3 I have lost more than fifteen pounds. | 3 | 1.1 |
| 19 | 0 I am no more worried about my health than usual. | 161 | 59.6 |
|  | 1 I am worried about physical problems like aches and pains. | 76 | 28.1 |
|  | 2 I am very worried about physical problems and it's hard to think much else. | 20 | 7.4 |
|  | 3 I am so worried about my physical problems that I can't think of anything. | 13 | 4.8 |

Tables 4.6a and 4.6b uses the Beck’s Depression Inventory to assess the level of depression amongst the respondents. The total score from each respondent was used to derive table 4.7.

**Table 4.7:** Prevalence of depression amongst respondents.

| *Variable* | *Frequency*<br>*(n=270)* | *Percentage*<br>*(%)* |
| --- | --- | --- |
| **No to mild depression (0 to 9)** | 157 | 58.1 |
| **Moderate depression (10 to 20)** | 84 | 31.1 |
| **Severe depression ( >20)** | 29 | 10.7 |
| **Min score amongst respondents: 0** |  |  |
| **Max score amongst respondents: 40** |  |  |
| **Mean score: 9.65 Std. dev: 8.41** |  |  |

The prevalence of mild depression amongst the respondents is 58.1%. The prevalence of moderate depression amongst the respondents is 31.1% while the prevalence of severe depression is 10.7%. The maximum score amongst the respondents was 40. The mean score was 9.65 + 8.41.

**Table 4.8:** Relationship between health risk behavior and sociodemographic characteristics of respondents.

| Sociodemography |  | Health risk Behaviour |  |  | Chi-square | df | p-value |
| --- | --- | --- | --- | --- | --- | --- | --- |
|  |  | No risk (%) | Low risk (%) | High risk (%) |  |  |  |
| Age | 14 years | 2 (10.5%) | 10 (52.6%) | 7 (38.8%) | 10.798 | 12 | 0.546 |
|  | 15 years | 4 (5.4%) | 48 (64.9%) | 22 (29.7%) |  |  |  |
|  | 16 years | 0 (0.0%) | 50 (66.7%) | 25 (33.3%) |  |  |  |
|  | 17 years | 2 (3.3%) | 36 (60.0%) | 22 (36.7%) |  |  |  |
|  | 18 years | 0 (0.0%) | 13 (76.5%) | 4 (23.5%) |  |  |  |
|  | 19 years | 0 (0.0%) | 9 (69.2%) | 4 (30.8%) |  |  |  |
|  | 20 years | 0 (0.0%) | 8 (66.7%) | 4 (33.3%) |  |  |  |
| Sex | Male | 1 (1.1%) | 48 (54.5%) | 39 (44.3%) | 8.962 | 2 | 0.011 |
|  | Female | 7 (3.8%) | 126 (69.2%) | 49 (26.9%) |  |  |  |
| Class | SSS1 | 3 (7.0%) | 29 (67.4%) | 11 (25.6%) | 12.017 | 4 | 0.017 |
|  | SSS2 | 1 (0.6%) | 105 (62.1%) | 63 (37.3%) |  |  |  |
|  | SSS3 | 4 (6.9%) | 40 (69.0%) | 14 (24.1%) |  |  |  |
| Are both parents alive? | Yes | 8 (3.1%) | 166 (64.8%) | 82 (32.0%) | 1.393 | 2 | 0.498 |
|  | No | 0 (0.0%) | 7 (53.8%) | 6 (46.2%) |  |  |  |
| Are you living with your parents | Yes | 7 (3.0%) | 146 (61.6%) | 84 (35.4%) | 6.366 | 2 | 0.041 |
|  | No | 1 (3.2%) | 26 (83.9%) | 4 (12.9%) |  |  |  |
| Are your parents living together | Yes | 8 (3.2%) | 157 (63.6%) | 82 (33.2%) | 1.193 | 2 | 0.551 |
|  | No | 0 (0.0%) | 16 (72.7%) | 6 (27.3%) |  |  |  |
| Are you in a love relationship | Yes | 3 (3.2%) | 63 (66.3%) | 29 (30.5%) | 0.217 | 2 | 0.897 |
|  | No | 5 (3.0%) | 105 (63.6%) | 55 (33.3%) |  |  |  |
| Do you use social media | Yes | 3 (1.6%) | 116 (60.7%) | 72 (37.7%) | 10.673 | 2 | 0.005 |
|  | No | 5 (6.4%) | 57 (73.1%) | 16 (20.5%) |  |  |  |
| Satisfied with school performance | Yes | 5 (3.2%) | 104 (66.2%) | 48 (30.6%) | 1.830 | 2 | 0.400 |
|  | No | 3 (3.1%) | 57 (58.2%) | 38 (38.8%) |  |  |  |
| Do you receive monthly allowances | Yes | 1 (1.6%) | 35 (56.5%) | 26 (41.9%) | 3.370 | 2 | 0.185 |
|  | No | 7 (3.4%) | 138 (66.7%) | 62 (30.0%) |  |  |  |

The prevalence of high risk behavior is higher in the males (44.3%) than in the females (26.9%). The prevalence of high risk behavior is highest in SSS2 students (37.3%) as compared to SSS1 (25.6%) and SSS3 (24.1%). The prevalence of high risk behavior is higher in respondents who are living with their parents (35.4%) than in respondents who are not living with their parents (12.9%). The prevalence of high risk behavior is higher in respondents who use social media (37.7%) than in respondents who don’t (20.5%). Therefore there is a significant relationship between gender, class, living with parents and the use of social media and health risk behavior (p<0.05). There is no significant relationship between age, both parents being alive, parents living together, love relationship and health risk behavior (p>0.05). The prevalence of high risk behavior is higher is respondents who are not satisfied with school performance (38.8%) than in respondents who are satisfied with school performance (30.5%). Also, the prevalence of high risk behavior is higher in respondents who receive monthly allowances (41.9%) than in respondents who don’t receive monthly allowances (30.0%). Though the relationship between school performance, monthly allowance and health risk behavior is not significant (p>0.05).

**Table 4.9:** Relationship between the level of depression and sociodemographic characteristics of respondents.

| Sociodemography |  | Level of depression |  |  | *Chi-square* | *df* | *p-value* |
| --- | --- | --- | --- | --- | --- | --- | --- |
|  |  | No to mild depression (%) | Moderate depression (%) | Severe depression (%) |  |  |  |
| **Age** | **14 years** | 7 (36.8%) | 7 (36.8%) | 5 (26.3%) | 24.656 | 12 | 0.017 |
|  | **15 years** | 51 (68.9%) | 19 (25.7%) | 4 (5.4%) |  |  |  |
|  | **16 years** | 38 (50.7%) | 25 (33.3%) | 12 (16.0%) |  |  |  |
|  | **17 years** | 39 (65.0%) | 18 (30.0%) | 3 (5.0%) |  |  |  |
|  | **18 years** | 11 (64.7%) | 6 (35.3%) | 0 (0.0%) |  |  |  |
|  | **19 years** | 7 (53.8%) | 5 (38.5%) | 1 (7.7%) |  |  |  |
|  | **20 years** | 4 (33.3%) | 4 (33.3%) | 4 (33.3%) |  |  |  |
| **Sex** | **Male** | 55 (62.5%) | 26 (29.5%) | 7 (8.0%) | 1.472 | 2 | 0.479 |
|  | **Female** | 102 (56.0%) | 58 (31.9%) | 22 (12.1%) |  |  |  |
| **Class** | **SSS1** | 19 (44.2%) | 18 (41.9%) | 6 (14.0%) | 5.220 | 4 | 0.265 |
|  | **SSS2** | 100 (59.2%) | 50 (29.6%) | 19 (11.2%) |  |  |  |
|  | **SSS3** | 38 (65.5%) | 16 (27.6%) | 4 (6.9%) |  |  |  |
| **Are both parents alive?** | **Yes** | 146 (57.0%) | 83 (32.4%) | 27 (10.5%) | 3.536 | 2 | 0.171 |
|  | **No** | 10 (76.9%) | 1 (7.7%) | 2 (15.4%) |  |  |  |
| **Are you living with your parents** | **Yes** | 136 (57.4%) | 75 (31.6%) | 26 (11.0%) | 0.174 | 2 | 0.916 |
|  | **No** | 19 (61.3%) | 9 (29.0%) | 3 (9.7%) |  |  |  |
| **Are your parents living together** | **Yes** | 148 (59.9%) | 75 (30.4%) | 24 (9.7%) | 5.824 | 2 | 0.054 |
|  | **No** | 8 (36.4%) | 9 (40.9%) | 5 (22.7%) |  |  |  |
| **Are you in a love relationship** | **Yes** | 47 (49.5%) | 37 (38.9%) | 11 (11.6%) | 5.705 | 2 | 0.058 |
|  | **No** | 105 (63.6%) | 42 (25.5%) | 18 (10.9%) |  |  |  |
| **Do you use social media** | **Yes** | 110 (57.6%) | 61 (31.9%) | 20 (10.5%) | 0.376 | 2 | 0.829 |
|  | **No** | 47 (60.3%) | 22 (28.2%) | 9 (11.5%) |  |  |  |
| **Satisfied with school performance** | **Yes** | 101 (64.3%) | 41 (26.1%) | 15 (9.6%) | 8.435 | 2 | 0.015 |
|  | **No** | 45 (45.9%) | 40 (40.8%) | 13 (13.3%) |  |  |  |
| **Do you receive monthly allowances** | **Yes** | 31 (50.0%) | 20 (32.3%) | 11 (17.7%) | 4.537 | 2 | 0.103 |
|  | **No** | 125 (60.4%) | 64 (30.9%) | 18 (8.7%) |  |  |  |

From the table above, the prevalence of severe depression is highest is respondents aged 20 years (33.3%), followed by those who are 14 years (26.3%) and those who are 16 years (16.0%). Though there is no clear cut increase or decrease in the prevalence of severe depression as the age increases, the relationship between age and depression is significant (p value < 0.05). The prevalence of severe depression is higher in the females (12.1%) than the males (8.0%). There is no significant relationship between gender and depression (p-value > 0.05). The prevalence of severe depression is higher in respondents who have less than two parents (15.4%) than in those with both parents alive (10.5%). Though the relationship between depression and that variable is not significant (p> 0.05). The prevalence of severe depression is higher in respondents whose parents are not living together (22.7%) than in respondents whose parents are living together (9.7%), though the relationship between depression and that variable is not significant (p>0.05). The prevalence of depression is higher in respondents who are not satisfied with school performance (13.3%), than in respondents who are satisfied with school performance (9.6%). There is a significant relationship between school performance and depression (p<0.05). There is no significant relationship between class, staying with parents, love relationship, social media and monthly allowance and depression (p>0.05).

**Table 4.10:** Relationship between the level of depression and health risk behavior of respondents.

|  |  | Level of depression |  |  | Chi-square | df | p-value |
| --- | --- | --- | --- | --- | --- | --- | --- |
|  |  | No to mild depression (%) | Moderate depression (%) | Severe depression (%) |  |  |  |
| **Health risk behavior** | **No risk** | 6 (75.0%) | 2 (25.0%) | 0 (0.0%) | 26.412 | 4 | 0.000 |
|  | **Low risk** | 118 (67.8%) | 45 (25.9%) | 11 (6.3%) |  |  |  |
|  | **High risk** | 33 (37.5%) | 37 (42.0%) | 18 (20.5%) |  |  |  |

From the table above, the prevalence of severe depression is highest in respondents with high risk behavior (20.5%), followed by respondents with low risk behavior (6.3%) and least in respondents with no risk behavior (0.0%). The relationship between health risk behavior and depression is highly significant (p<0.001).

**Table 4.11:** To determine the health risk behavior of respondents associated with depression.

| Health Risk Behaviors |  | Level of depression |  |  | Chi-square | df | p-value |
| --- | --- | --- | --- | --- | --- | --- | --- |
|  |  | No to mild depression (%) | Moderate depression (%) | Severe depression (%) |  |  |  |
| Have you ever tried smoking? | Yes | 9 (42.9%) | 8 (38.1%) | 4 (19.0%) | 2.736 | 2 | 0.255 |
|  | No | 148 (59.4%) | 76 (30.5%) | 25 (10.0%) |  |  |  |
| Have you ever taken alcoholic drinks? | Yes | 96 (53.9%) | 61 (34.3%) | 21 (11.8%) | 3.815 | 2 | 0.148 |
|  | No | 61 (66.3%) | 23 (20.5%) | 8 (8.7%) |  |  |  |
| Have you used marijuana or weed? | Yes | 5 (74.1%) | 1 (14.3%) | 1 (14.3%) | 0.956 | 2 | 0.620 |
|  | No | 152 (57.8%) | 83 (31.6%) | 28 (10.6%) |  |  |  |
| Are you sexually active? | Yes | 9 (52.9%) | 5 (29.4%) | 3 (17.6%) | 0.907 | 2 | 0.635 |
|  | No | 148 (58.5%) | 79 (31.2%) | 26 (10.3%) |  |  |  |
| Have you been pregnant or gotten someone pregnant? | Yes | 1 (25.0%) | 2 (50.0%) | 1 (25.0%) | 2.001 | 2 | 0.368 |
|  | No | 156 (58.6%) | 82 (30.8%) | 28 (10.5%) |  |  |  |
| Do you think you are physically unfit? | Yes | 21 (48.8%) | 13 (30.2%) | 9 (20.9%) | 5.719 | 2 | 0.057 |
|  | No | 136 (59.9%) | 71 (31.3%) | 20 (8.8%) |  |  |  |
| Do you want to lose or gain weight? | Yes | 124 (59.6%) | 63 (30.3%) | 21 (10.1%) | 0.883 | 2 | 0.643 |
|  | No | 33 (53.2%) | 21 (33.9%) | 8 (12.9%) |  |  |  |
| Have you carried a weapon (gun or knife) to school? | Yes | 3 (50.0%) | 2 (33.3%) | 1 (16.7%) | 0.280 | 2 | 0.869 |
|  | No | 154 (58.3%) | 82 (31.1%) | 28 (10.6%) |  |  |  |
| Have you fought in the past 6 months? | Yes | 28 (45.9%) | 22 (36.1%) | 11 (18.0%) | 6.556 | 2 | 0.038 |
|  | No | 129 (61.7%) | 62 (29.7%) | 18 (8.6%) |  |  |  |
| Have you ever thought of killing yourself? | Yes | 13 (23.6%) | 29 (52.7%) | 13 (23.6%) | 32.215 | 2 | 0.000 |
|  | No | 144 (67.0%) | 55 (25.6%) | 16 (7.4%) |  |  |  |

From the table above, the major health risk behaviors associated with depression are fighting and contemplation of suicide. The prevalence of severe depression is higher in respondents who have fought in the past 6 months (18.0%), than in those who haven’t (8.6%). The prevalence of severe depression is higher is respondents who have contemplated suicide (23.6%), than in respondents who haven’t (7.4%). The relationship between fighting and depression is significant (p<0.05), while the relationship between suicidal thoughts and depression is highly significant (p<0.001).

## CHAPTER FIVE

### DISCUSSION

The study aimed to find out the prevalence of depression among senior secondary school students in Enugu Metropolis. The sample was made up of 270 students drawn from both public and private secondary schools within Enugu Metropolis. Hard copies of the questionnaire was shared and a 100% response achieved.

The age distribution was between 14-20, with the average at 16.26 and a standard deviation of 1.46 years. Most of the respondents were in the age range of 15-17 and were from the SSS2 class. About 32.6% (88 students) were male and 67.4% (182) were female. Almost all the respondents were of the Igbo tribe, from the South Eastern part of the country and were also Christians. A majority of the respondent’s parents’ occupation was business.

The results of this study showed that concerning the levels of depression among the studied population that, the prevalence of mild depression was 58.1% (157 students), the prevalence of moderate depression was 31.1% (84 students) and the prevalence of severe depression was 10.7% (29 students).

An important objective of this study was to determine the socio demographic factors associated with depression among the sample population and about this, the study found that the prevalence of severe depression was highest in respondents aged 20 years (33%), followed by those who are 14 years (26.3%) and those who are 16 years (16.0%). Although there is no clear cut increase or decrease in the prevalence of severe depression as the age increases or decreases, the relationship between age and depression is significant (p value<0.05).

This finding is in keeping with a study done by Oderinde et al ^[6]^ which found out that proportion of older adolescents in the age range 16-19 who were found to be depressed was higher than the proportion of younger adolescents in the age range 10-12 years who were found to be depressed. This study also found that the prevalence of severe depression was higher in the females (12.1%) than the males (8.0%), which is in agreement with several studies done around this topic. Gender was named among the most significant predictors of depression in a study done by Al-Kaabi et al.^[14]^ The prevalence of severe depression was also higher in respondents who have less than two parents (15.4%) than in those with both parents alive (10.5%) although the relationship between depression and this variable is not significant (p value>0.05). The reason for this observation most likely to be that such students are often deprived of a secure and loving relationship with their parents and these are the protective factors that reduce the rate of emotional disorders among adolescents.

The prevalence of severe depression was higher in respondents whose parents are not living together (22.7%) than in those whose parents are living together (9.7%), though the relationship between depression and this variable is not significant (p value>0.05). This could be owing to the fact the parents are divorced or separated because of one reason or the other and the children end up with little or a non-existent relationship with their parents and this in turn has some psychosocial effects on them. The prevalence of depression was found to also be higher in respondents who were not satisfied with school performance (13.3%) than in those who were satisfied with their school performance (9.6%). There is a significant relationship between school performance and depression (p value<0.05)

The study found that there was no significant relationship between class, staying with parents, love relationship, social media use and monthly allowance and depression (p value>0.05)

Concerning health risk behaviours and its association with depression, it is important to note that this study found a highly significant relationship between depression and health risk behaviours (p<0.001) The prevalence of severe depression was highest in respondents with high risk behaviours (20.5%), followed by those with low risk behaviour (6.3%).

In line with one of the objectives of this study which was to determine some health risk behaviours associated with depression, this study found that the major health risk behaviours associated with depression were fighting and contemplation of suicide.

The prevalence of severe depression was higher in respondents who had fought in the past six months (18.0%), than in those who hadn’t (8.6%). the prevalence of severe depression was also higher in those who had contemplated suicide (23.6%), than in those who hadn’t (7.4%). The relationship between fighting and depression is significant (p<0.05) and the relationship between suicidal thoughts and depression is highly significant (p<0.001).

This study was carried out during the covid-19 pandemic and so the limitation we encountered was with respect to administering the questionnaire to the study population. This was because school resumption dates following lifting of the lockdown was variable and collecting data especially from the SSS 3 students posed a big problem because majority of them were not in school, data gotten from this class was collected prior to the lockdown.

In line with this study, we recommend gross sensitization of parents, teachers, school social workers and school nurses on the signs and symptoms of adolescent depression, this is to facilitate early diagnosis and prompt institution of therapy. Screening programs for adolescent depression should be included in to the primary health care scheme to ensure inclusion across all socio economic classes and this should made as frequent as possible.

## CHAPTER SIX

### CONCLUSION AND RECOMMENDATIONS

This study performed on the prevalence of depression among senior secondary schools in Enugu metropolis showed a significant level of prevalence of depression in the students in the study population.. The study showed that depression was higher in the female gender, those with less than two parents, students not living with their parents and those who were not satisfied with their academic performance. A significant relationship was found between depression, dissatisfaction with academic performance and health risk behaviors. The health risk behaviors associated with depression were fighting and suicidal tendencies. However, there was no significant relationship between age, both parents being alive, parents living together, love relationships and health risk behaviors.

The first limitation encountered in this study was difficulty in administering the printed questionnaire to the students in the study population due to school closure in the wake of the Corona virus pandemic and subsequent lockdown.

Another one was that the study was done with a cross sectional study design so the changes in prevalence of depression over time could not be ascertained.

In line with our findings in this study, we are recommending as follows:

1. Awareness of the possibility of depression among adolescents in secondary schools should be carried out through adequate sensitization of the general public through mass media and publications.
2. Teachers, school social workers, nurses and parents should be educated about the symptoms of depression to assist in early recognition and institution of therapy.
3. Provision of mental health awareness programs and outreach in secondary schools.
4. Screening for Adolescent depression in primary health care should be provided and frequent.
5. 3. Counseling, support and guidance services should be made available to students with depressive symptoms.
6. Use of standardized screening tools to improve identification of students with depression
7. Following diagnosis of any level of depression, appropriate support and therapy system should be instituted and patient followed up closely.
8. Discouraging high risk behaviors like smoking, use of hard drugs, risky sexual behaviors among secondary school students.
9. Studies conducted to monitor the changes in prevalence of depression among secondary school students over time should be encouraged and funded.

## Data Availability

There are no restrictions to data availability

## ACKNOWLEDGEMENTS

Our heartfelt gratitude goes to our parents encouraged us financially and in numerous other ways. We also wish to acknowledge our wonderful supervisor Prof. E.N. Aguwa for his patience, dedication and support throughout this project and also for pushing us consistently towards excellence and perfection. We would also like to acknowledge our dear friends and classmates for their support towards our success. Our special thanks goes to Dr. Ifeoma Obionu for her advice, patience and direction towards making this project a success.

## REFERENCES

1. Keiling C, Baker-Henninghan H, Belfer M, Conti G, Ertem I, Omigbodun O. Child and adolescent mental health worldwide: Evidence for action. 2011;378(9801)

2. Chinawa J, Manyike P, Obu H, Aronu A, Odutola O, Chinawa A. Depression among adolescents attending secondary school in South East Nigeria. Annals of African Medicine. 2015;14(1):46–51

3. WHO adolescent Mental Health. Available from https://www.who.int/news-room/fact-sheets/detail/adolescent-mental-health (Accessed on 13 January 2020)

4. Lancet Global Health. Available from https://www.thelancet.com/journals/langlo/article/PIIS2214-109X(18)30303-6/fulltext (Accessed on 13 January 2020)

5. Population Health Metrics. Available from https://pophealthmetrics.biomedcentral.com/articles/10.1186/s12963-016-0084-2 (Accessed on 14 January 2020)

6. Oderinde K, Dada M, Ogun O, Awunor N, Kundi B, Ahmed H, Tsuung A, Tanko S, Yusuff A. Prevalence and Predictors of Depression among Adolescents in IdoEkiti, South West Nigeria. International Journal of Clinical Medicine, 2018; 9:187–202.

7 . Emslie G, Mayes T. Depression in children and adolescents: A guide to diagnosis and treatment. CNS Drugs. 1999; 11: 181–189.

8. Omigbodun O, Esan O, Bakare K, et al. Depression and Suicidal Symptoms among Adolescents in Rural South Western Nigeria. 16th World Congress of the International Association for Child and Adolescent Psychiatry and Allied Professions (IACAPAP), Berlin. 2004;22–26.

9. Omigbodun O, Bakare K, Yusuf B. Traumatic Events and Depressive Symptoms among Youths in South West, Nigeria: A Qualitative Analysis. International Journal of Adolescent Medical Health. 2008; 20:243–253.

10. Fatiregun A, Kumapayi T. Prevalence and Correlates of Depressive Symptoms among School Adolescents in a Rural District in South West. Nigeria.Journal of Adolescence. 2014;37:197–203.

11. Nagendra K, Sangay D, Gouli C, Kalappanavar N. Prevalence and Association of Depression and Suicidal Tendency among Adolescent Students in Davangere, India. International Journal of Biomedical and Advance Research. 2012;3:714–918.

12. Bahls S. Epidemiology of Depressive Symptoms in Adolescents of a Public School in Curitiba, Brazil. Brazilian Journal of Psychiatry. 2002; 24:63–67.

13. Frank E, Carpenter L, Kupfer D. Sex Differences in Recurrent Depression: Are There Any That Are Significant? The American Journal of Psychiatry. 1988; 145:41–45.

14. Al-Kaabi N, Selim NAA, Singh R, Almadahki H, Salem M. Prevalence and Determinants of Depression among Qatari Adolescents in Secondary Schools. Fam Med MedSci Res 2017;6: 219.

15. Derdikman-Eiron R, Indredavik MS, Bratberg GH, Taraldsen G, Bakken IJ, et al. Gender differences in subjective well being, self esteem and psychosocial functioning in adolescents with symptoms of anxiety and depression: findings from the Nord Trondelag Health Study. Scand J Psychol. 2011;52: 261–267.

16. Martin LA, Neighbors HW, Griffith DM. The experience of symptoms of depression in men vs women: analysis of the National Comorbidity Survey Replication. JAMA Psy. 2013; 70: 1100–6.

17. Schoenfelder EN, Sandler IN, Wolchik S, MacKinnon D. Quality of Social Relationships and the Development of Depression in Parentally Bereaved Youth. J Youth Adol. 2011;40: 85–96.

18. Kessle R, McGonagle K, Swartz M, et al. Sex and Depression in the National Comorbidity Survey I: Lifetime Prevalence, Chronicity and Recurrence. Journal of Affective Disorders. 1993; 29:85–96.

19. Abdel-Azis M, Abdel-Hady N, Elimissiry S, El-Rasheed A, Sabry W. Prevalence of Depression in a Sample of Egyptian Secondary School Female Students. Egyptian Journal of Psychiatry. 2013; 1–12.

20. Adams, D.B. Parental Death and Teenage Depression. American Journal of Psychiatry. 2009;166: 786–794.

21. Slavich G, Monroe S, Gotlib I. Early Parental Loss and Depression History: Associations with Recent Life Stress in Major Depressive Disorder. Journal of Psychiatric Research. 2011; 45: 1146–1152.

22. JAMA and Archives Journals (2011) Grief Reactions Subside in Most Children and Teens Whose Parents Die Suddenly but May Persist or Increase in Some Cases. (Internet). Science Daily

23. GBD 2017 Disease and Injury Incidence and Prevalence Collaborators. Global, regional, and national incidence, prevalence, and years lived with disability for 354 diseases and injuries for 195 countries and territories, 1990–2017: a systematic analysis for the Global Burden of Disease Study 2017. The Lancet. 2018. DOI.

24. Wang, et al. Use of mental health services for anxiety, mood, and substance disorders in 17 countries in the WHO world mental health surveys. The Lancet. 2007; 370(9590):841–50.

25. American Psychiatric Association: Diagnostic and Statistical Manual of Mental Disorders, Fifth Edition: DSM-5, American Psychiatric Publishing, 2013.

26. National Alliance on Mental Illness: “Criteria for Major Depressive Episode: DSM-5,” “Psychotic.”Parker, G. The American Journal of Psychiatry, September 2002.

27. Morbidity and Mortality Weekly Report. The American Journal of Psychiatry. May 2012.

28. Severus, E. Anxiety and Depression Association of America: “Depression. International Journal of Bipolar Disorders. 2013.

29. Psychiatry MMC: “Atypical Depression, PubMed. Diagnostic and Statistical Manual Disorders (DSM-5), Fifth Edition. National Institute of Mental Health.

30. Enugu state. Available from https://en.m.wikipedia.org/wiki/Enugu_state (Accessed on 26th January, 2020.)

31. Becks Depression Inventory (BDI-I).

32. Pierre V, Caroline L, Bruno V. Is depression associated with health risk-related behaviour clusters in adults?. European Journal of Public Health. 2009; 19(6)618–624.

33. Depression and risk behaviours in Adolescents. Available from https://www.ncbi.nlm.nih.gov/pubmed/24707767 (Accessed on the 18 January 2020).

34. Pailing AN, Reniers RLEP. Depressive and socially anxious symptoms, psychosocial maturity, and risk perception: Associations with risk-taking behaviour. PLoS ONE. 2018;13(8)

35. Youth Risk Behaviour Surveillance System (YRBSS) Questionnaire, 1991-2019.

